# Assessment of a zero-shot large language model in measuring documented goals-of-care discussions

**DOI:** 10.1101/2025.05.23.25328115

**Authors:** Robert Y. Lee, Kevin S. Li, James Sibley, Trevor Cohen, William B. Lober, Danae G. Dotolo, Erin K. Kross

## Abstract

**Context:** Goals-of-care (GOC) discussions and their documentation are important process measures in palliative care. However, existing natural language processing (NLP) models for identifying such documentation require costly task-specific training data. Large language models (LLMs) hold promise for measuring such constructs with fewer or no task-specific training data.

**Objective:** To evaluate the performance of a publicly available LLM with no task-specific training data (zero-shot prompting) for identifying documented GOC discussions.

**Methods:** We compared performance of two NLP models in identifying documented GOC discussions: Llama 3.3 using zero-shot prompting; and, a task-specific BERT (Bidirectional Encoder Representations from Transformers)-based model trained on 4,642 manually annotated notes. We tested both models on records from a series of clinical trials enrolling adult patients with chronic life-limiting illness hospitalized over 2018-2023. We evaluated the area under the receiver operating characteristic curve (AUC), area under the precision-recall curve (AUPRC), and maximal F_1_ score, for both note-level and patient-level classification over a 30-day period.

**Results:** In our text corpora, GOC documentation represented <1% of text and was found in 7.3-9.9% of notes for 23-37% of patients. In a 617-patient held-out test set, Llama 3.3 (zero-shot) and BERT (task-specific, trained) exhibited comparable performance in identifying GOC documentation (Llama 3.3: AUC 0.979, AUPRC 0.873, and F_1_ 0.83; BERT: AUC 0.981, AUPRC 0.874, and F_1_ 0.83).

**Conclusion:** A zero-shot large language model with no task-specific training performed similarly to a task-specific trained BERT model in identifying documented goals-of-care discussions. This demonstrates the promise of LLMs in measuring novel clinical research outcomes.

**KEY MESSAGE:** This article reports the performance of a publicly available large language model with no task-specific training data in measuring the occurrence of documented goals-of-care discussions from electronic health records. The study demonstrates that newer large language AI models may allow investigators to measure novel outcomes without requiring costly training data.

## Introduction

Advances in natural language processing (NLP) hold great promise for allowing researchers to measure previously inaccessible constructs from unstructured electronic health records (EHR). Because many important clinical constructs in EHRs are linguistically complex and difficult to capture with rule-based or keyword searches,^1^ researchers have used machine learning (ML) models trained on manually curated and labeled datasets—a process called *supervised learning*—to extract complex constructs from EHR text.^2^ However, acquiring and labeling training datasets for supervised learning can be prohibitively expensive, especially for rare constructs. This problem, which has been termed the “annotation bottleneck,” is an important barrier for the use of supervised learning in clinical applications of NLP.^2^

Many recent advances in NLP have been driven by large language models (LLMs), a newer class of ML models pre-trained on vast and diverse corpora of unlabeled text. Through their pre-training, LLMs acquire a broad representation of linguistic structures and relationships that allow them to perform a wide range of NLP tasks with little or no task-specific training data.^3^ Recent generations of LLMs have gained widespread attention through their deployment as artificial intelligence (AI) chatbots, which at least imitate human-like knowledge in many areas.^4^ As such, there has been growing interest in using LLMs to measure linguistic constructs without the need for large amounts of training data. In these approaches, LLMs are prompted to classify text with minimal or no task-specific training data (referred to as *few-shot* and *zero-shot learning*, respectively), instead relying on careful prompting in conjunction with the model’s “out-of-the-box” pre-training to apply previously learned patterns to the task at hand (**Table 1**).^5^

**Table 1.** Comparison of supervised, few-shot, and zero-shot machine learning.

| Learning approach | Description <sup>a</sup> | Examples and analogies |
| --- | --- | --- |
| **Supervised learning** | Model is trained ("fine-tuned") on a labeled dataset, mapping input data to the correct output by minimizing error between predicted and actual labels. | ▪ ML model to distinguish between dogs from cats trained on 1,000 pictures of dogs and 1,000 pictures of cats, each labeled accordingly<br>▪ ML model to identify documented GOC discussions trained on 5,000 EHR notes, of which 300 are labeled as containing GOC discussions |
| **Few-shot learning** | Pre-trained model is fine-tuned to, or prompted with, a small number of labeled examples, relying on the model's pre-training to apply previously learned patterns to a new task. Examples need not cover full range of variability or noise expected in the target task, as the model leverages prior learning to generalize effectively. | ▪ ML model with generalized pre-training fine-tuned on just 6 pictures of dogs and cats to distinguish between them<br>▪ A child learning to distinguish airplanes from helicopters after being shown 5 examples from a picture book<br>▪ LLM prompted to identify documented GOC discussions after being given a small number of examples |
| **Zero-shot prompting** | Model is prompted to make predictions without any task-specific examples, relying entirely on the model's pre-training to generalize to the new task. | ▪ Vision-language model with generalized pre-training asked to distinguish between dogs and cats, with no fine-tuning or task-specific examples<br>▪ Resident physician asked to characterize an unusual chest x-ray finding<br>▪ LLM prompted to identify documented GOC discussions by definition alone, with no provided examples |

Abbreviations: ML, machine learning; LLM, large language model; GOC, goals of care.
<sup>a</sup> In machine learning, *pre-training* refers to the initial process of training a model on a large, general dataset to learn patterns, representations and features that are broadly applicable across tasks. Pre-training is often performed by the model's creators. Whereas, *fine-tuning* refers to subsequent adjustment of a pre-trained model on a smaller, task-specific dataset, often performed by those adapting the model for a particular use.

Goals-of-care discussions are an important process measure in palliative care research, and likely mediate the delivery of patient-centered, goal-concordant care.^6–8^ However, timely and high-quality assessment of patients’ goals of care remains a significant shortcoming in health systems.^9–11^ Additionally, because goals-of-care discussions are frequently documented in unstructured text such as progress notes or other inconspicuous locations, documentation of historical discussions can be difficult for clinicians to unearth during clinical care.^12–14^ This has led to interest in using NLP to measure goals-of-care discussions.^15–20^ To date, the use of NLP to identify this linguistically complex outcome has facilitated at least two clinical trials;^19–22^ and, while current clinical EHRs rely on either search-based approaches or the use of semi-structured documentation,^12,23^ real-time ability for clinicians to efficiently locate goals-of-care documentation seems likely to improve delivery of goal-concordant care.^24^

Our research group has studied the use of NLP to identify EHR-documented goals-of-care discussions for patients with serious illness. In our previous work, a supervised learning model based on BERT (Bidirectional Encoder Representations from Transformers)^25,26^ demonstrated promising performance in identifying goals-of-care documentation for hospitalized patients, with an area under the receiver operating characteristic curve [AUC] of 0.962 and area under the precision-recall curve [AUPRC] of 0.824 for adjudicating individual notes.^19^ However, acquiring the task-specific training and validation data for this model required hundreds of human abstractor-hours,^19^ limiting our ability to adapt this model toward measuring other constructs or conducting larger trials.

In a recent report, a zero-shot LLM was used to identify advance care planning and goals of care documentation for outpatient oncology patients.^18,27^ To our knowledge, no study has yet evaluated the use of zero-shot LLMs to identify goals-of-care discussions in a general population of seriously ill hospitalized patients against manually adjudicated standards. To explore the potential of modern LLMs for current and future research tasks, we evaluated the performance of a publicly accessible LLM using zero-shot prompting to identify documented goals-of-care discussions for hospitalized patients with serious illness.

## Methods

This diagnostic study evaluated the zero-shot performance of Llama 3.3 (Meta AI, ai.meta.com), a publicly available pre-trained general-purpose LLM, in predicting the presence of documented goals-of-care discussions in EHR notes of hospitalized patients with serious illness. We compared Llama predictions against gold-standard results from manual abstraction, and also compared Llama performance against the benchmark of a previous BERT-based model that was fitted using supervised ML to manually-abstracted training data.^19^ All study procedures were approved by the University of Washington Human Subjects Division Institutional Review Board (STUDY00011002).

### Data sources

There are three datasets used in this trial (**Table 2**): (1) a pilot set, comprising 4,642 EHR notes from 150 patients enrolled in a pilot trial of a patient- and clinician-facing communication-priming intervention;^19,28^ (2) the Trial 1 test set, comprising 2,974 EHR notes from a sample of 160 of 2,512 patients enrolled in a pragmatic trial of a clinician-facing-only communication-priming intervention;^21,29^ and, (3) the Trial 2 held-out test set, comprising 11,574 EHR notes from 617 patients enrolled in a comparative-effectiveness trial of two communication priming interventions.^29^ The detailed inclusion criteria for patients in each dataset have been previously reported.^19,21,28,29^ Briefly, all patients were hospitalized at any of 2-3 study hospitals in the UW Medicine health system, Seattle, WA; were of advanced age (≥ 80 years); or, were of age 55 years or older with one or more chronic life-limiting illnesses as defined by diagnosis codes. Each dataset consisted of all notes for the sampled patients collected over similar time frames that were authored by attending and trainee physicians, advance practice providers, and subinterns. Patients in the Trial 1 test set were randomly selected from all enrolled participants on the date of an interim analysis in February 2021, with intentional enrichment for patients with Alzheimer disease and related dementias (ADRD; 80/160 [50%]) to support an interim subgroup analysis in the parent trial. (The Trial 1 test set is slightly larger here than in previous reports due to the inclusion of notes preceding parent trial randomization and notes from one patient whose records were inadvertently sampled from the wrong index admission.) The interventions tested in all three trials (variants of the Jumpstart Guide) did not alter existing clinician EHR documentation processes or templates for documentation of goals-of-care discussions.^21,28,29^

**Table 2.** Datasets used in this study.

|  | Pilot set | Trial 1 test set | Trial 2 held-out test set |
| --- | --- | --- | --- |
| **Description of patients, and enrollment procedure** | All 150 patients enrolled in a 2-hospital pilot trial with informed consent over Nov. 2018 to Feb. 2020 <sup>28</sup> | Sample of 160 patients enrolled in a 2,512-person 3-hospital pragmatic trial under a waiver of informed consent, enriched for ADRD (80 of 160 [50%]), over Apr. 2020 to Mar. 2021 <sup>21,29</sup> | All 617 patients enrolled in a 3-hospital comparative-effectiveness trial with informed consent over Jul. 2021 to Nov. 2023 <sup>29</sup> (trial results not yet published) |
| **Description of EHR notes** | 4,642 notes from index admission to discharge | 2,974 notes from index admission to 30 days post-randomization | 11,574 notes from randomization to 30 days post-randomization |
| **Adjudication of ground truth** | Manual whole-chart abstraction by human reviewers, with regular quality assurance applied at the passage level | Manual whole-chart abstraction by human reviewers, with regular quality assurance applied at the passage level | BERT NLP-screened human abstraction of passages scoring over 98.5<sup>th</sup> percentile, <sup>a</sup> with principal investigator co-review of all positive abstractions by patient |
| **Prevalence of GOC discussions** | 0.2% of BERT segments; 340 / 4,642 (7.3%) notes; 34 / 150 (23%) patients | 0.4% of BERT segments; 295 / 2,974 (9.9%) notes; 59 / 160 (37%) patients | 304 / 2,136 notes in case-control sample; <sup>b</sup> 163 / 617 (26%) patients |
| **Role of dataset for each NLP model <sup>c</sup>** |  |  |  |
| **Llama 3.3 LLM**<br>(zero-shot prompt) | Development | Testing | Held-out testing |
| **BERT**<br>(supervised ML) | Training and validation | Testing | Held-out testing |

Abbreviations: ADRD, Alzheimer disease and related dementias; EHR, electronic health record; BERT, Bidirectional Encoder Representations from Transformers; GOC, goals of care; NLP, natural language processing; LLM, large language model; ML, machine learning.
<sup>a</sup> In the Trial 1 test set, this screening threshold corresponded with 99.3% note-level sensitivity (95%CI 97.6%, 99.9%) and 100% patient-level sensitivity (one-sided 97.5%CI 97.7%, 100%).
<sup>b</sup> Case-control sample of notes in the Trial 2 held-out test: cases, 304 notes with human-confirmed GOC content; controls, up to three randomly sampled GOC-negative notes per patient. Prevalence in sample (14%) is not expected to represent source data.
<sup>c</sup> In supervised machine learning, “training” refers to fitting a model to labeled data; “validation” refers to tuning model hyperparameters during development; and, “testing” refers to evaluating the performance of the final model after all training and development is complete. A “held-out test set” is a special test set that is kept completely separate from all model development, minimizing possibility of indirect leakage or bias. Because zero-shot prompting does not fit a model to any labeled data, the pilot set is referred to as a “development” set for the Llama model.

By definition, the zero-shot Llama 3.3 LLM had no task-specific training dataset, and all LLM prompt engineering and hyperparameter selection was conducted using notes drawn from the pilot set. The Llama model was tested on both the Trial 1 test set and the Trial 2 held-out test set. The reference BERT-based model was trained and validated on the pilot set, and subsequently evaluated on the Trial 1 test set as previously reported.^19^ For this study, the same model was also tested on the Trial 2 held-out test set.

### Natural language processing

Llama 3.3 is a 70-billion-parameter generative LLM with a 128,000-token context length, developed and pre-trained by Meta AI on an undisclosed collection of public online data. (*Tokens* are an NLP term for words and subwords that form the smallest unit of NLP analysis; the *context length* of a model defines the largest text passage it can analyze at once.) Most EHR notes may be analyzed in their entirety within Llama 3.3’s context length; notes exceeding this context length were partitioned evenly and assigned the highest predicted probability of their constituent partitions. Notes were preprocessed using regular expressions to remove common headers, footers, allergy and medication sections, and a palliative care quality improvement checklist consisting only of discrete data. To predict the probability of documented goals-of-care discussions in each candidate note or partition, we prompted a newly initialized Llama 3.3 model with our operational definition and the candidate text (**Appendix 1**). We then examined the raw scores (logits) underlying the model’s generated response, which were transformed into probabilities that reflect the model’s confidence in predicting an affirmative (“Yes”) vs. negative (“No”) response.

The reference BERT-based model for identifying documented goals-of-care discussions^19^ is an instance of BioClinicalBERT^26^ that was trained by our team on labeled notes from the pilot set (**Table 2**). BioClinicalBERT is a publicly-available customization of BERT_BASE_ (Bidirectional Encoder Representations from Transformers—base model),^25^ a 110-million-parameter model with a 512-token context length developed and pre-trained by Google Research (google.com) on the full text of English Wikipedia and BookCorpus (a dataset of 11,000 unpublished books).^30^ BioClinicalBERT was further pre-trained by Alsentzer *et al* on PubMed abstracts, PubMed Central manuscripts, and EHR notes from the MIMIC III database.^26,31^ Notes were partitioned into *BERT passages* of ≤ 512 BERT tokens over common whitespace patterns; the BERT-predicted probability for each note was defined by the highest predicted probability of its constituent passages.

Llama 3.3 and other recent Llama models have been released by Meta AI under a licensing agreement and use policy that allows for free non-commercial use by the public. To ensure privacy and prevent model developers from training on protected health information, we ran Llama locally on a secure in-house server equipped with a 16-core AMD Epyc 9124 processor (Advanced Micro Devices, amd.com), a single Nvidia RTX A6000 graphics processing unit (GPU) (Nvidia Corporation, nvidia.com), and 48 gigabytes of video random-access memory (VRAM). Both BioClinicalBERT and the original BERT family of models have been made open-source and publicly available by their respective developers.^32,33^

### Adjudication of ground truth

For this study, goals-of-care discussions were defined as discussion of the overarching aims of medical care for a patient^34^ that extended beyond routine code status discussions or standalone citations of past advance care planning documents. This pragmatic definition was designed to capture typical documentation of discussions by both specialty palliative care and other clinicians, while excluding routine admission code status discussions or automated listings of advance care planning documents on file.^21,35^ The definition was operationalized using a medical record abstraction manual that has been previously published.^19^

Ground-truth for BERT passage- and note-level presence or absence of EHR-documented goals-of-care discussions was determined for all notes in the pilot set and Trial 1 test set using manual human chart abstraction, as previously described.^19^ During these abstraction efforts, abstractors met regularly as a group to discuss their findings, and instances of disagreement were resolved by consensus. Quality assurance practices, such as co-review of passages by senior research coordinators and investigators, were applied at the passage level.

The Trial 2 held-out test set, which included 617 patients—substantially more than the pilot (150 patients) and Trial 1 (160 patients) test sets—was adjudicated for presence and timing of first human-confirmed documented goals-of-care discussion within 30 days of randomization using BERT NLP-screened human abstraction. In this approach, the BERT model, which was trained on the pilot set and previously tested on the Trial 1 test set, was applied to the Trial 2 held-out test set using a highly-permissive (i.e., sensitive) screening threshold to select passages for human adjudication for goals-of-care content. This approach enabled measurement of a much larger number of patient-level outcomes than would be feasible in a manually-abstracted validation sample. For adjudicating the held-out test set, the screening threshold was set to the 98.5^th^ percentile of BERT-predicted passage-level probabilities for goals-of-care content, resulting in 8,952 of 559,596 BERT passages (1.6%) screening positive; 3,718 of 11,574 notes (32%) contained at least one screen-positive passage, and 533 of 617 patients (86%) had at least one screen-positive passage. Evaluation of the selected BERT screening threshold in the Trial 1 test set yielded 99.3% note-level sensitivity (95%CI 97.6%, 99.9%) and 100% patient-level sensitivity (one-sided 97.5%CI 97.7%, 100%).^19^ Because not all BERT-positive notes in the Trial 2 held-out test set were adjudicated by humans, note-level performance in the Trial 2 held-out test set was evaluated using a 2,136-note case-control sample of all notes containing human-confirmed goals-of-care content (304 notes) and up to 3 randomly selected notes per patient without goals-of-care content (i.e., with BERT score less than 98.5^th^ percentile; 1,832 notes). In addition to the same quality assurance practices applied during abstraction of the pilot and Trial 1 test sets, Trial 2 records coded as positive or possibly-positive by abstractors were further co-reviewed by a single clinician (E.K.) in the context of the patient’s chart to ensure consistency of the outcome measure. Patient-level performance in the Trial 2 held-out test set was evaluated by comparing NLP predictions for all 11,574 notes against patient-level results of NLP-screened human abstraction for all 617 patients.

### Statistical analysis

We evaluated the performance of each language model using receiver operating characteristic (ROC) curve analysis, which compares sensitivity and specificity across all discrimination thresholds; precision-recall (PR) analysis, which compares positive predictive value (PPV; or, precision) and sensitivity (recall) across all discrimination thresholds;^36^ and, evaluation of the F_1_ score, a metric commonly used to describe the predictive performance of information retrieval systems that is defined as the harmonic mean of PPV and sensitivity (range, 0 to 1). ROC and PR analyses were evaluated both graphically and also by their respective areas-under-the-curve, with AUC indicating area under the ROC curve, and AUPRC indicating area under the precision-recall curve. As F_1_ is a threshold-dependent measure, we reported the highest observed F_1_ score across all discrimination thresholds. In all analyses, patient-level probability was defined as the maximal NLP-predicted probability of all constituent notes or passages; and, patient-level ground truth was defined as the union of ground truth for all constituent notes. Statistical analyses were performed using Stata/MP, version 19.0 (StataCorp LLC, stata.com). AUC and AUPRC were calculated using cubic splines.

## Results

The composition, inclusion criteria, and adjudication of ground truth for each dataset are described in **Table 2**. In each dataset, there were between 295 and 340 notes containing goals-of-care discussions, and between 1,832 and 4,302 notes without goals-of-care discussions. In the Trial 1 test set, 59 of 160 patients (37%) had at least one goals-of-care discussion between index admission and 30 days post-randomization. In the Trial 2 held-out test set, 163 of 617 patients (26%) had at least one goals-of-care discussion between index admission and 30 days post-randomization.

In comparing Llama 3.3 and BERT NLP predictions against manual or NLP-screened manual abstraction in the Trial 1 test set and Trial 2 held-out test set, the zero-shot Llama model consistently demonstrated highly comparable performance to the reference BERT model that had been trained on the pilot set. **Figure 1** shows ROC and precision-recall curves for note-(panel A) and patient-level (panel B) classifiers across models (Llama vs. BERT) and datasets, and **Table 3** shows areas under the ROC and precision-recall curves as well as maximum observed F_1_ scores for each model and dataset. In the Trial 2 held-out test set, the area under the ROC curve for note-level analysis was 0.979 for Llama, and 0.981 for BERT; and, the area under the precision-recall curve for note-level analysis was 0.873 for Llama, and 0.874 for BERT. The maximum observed note-level F_1_ score in the held-out test set was 0.83 for both Llama and BERT.

**Table 3.** Performance metrics of zero-shot Llama 3.3 vs. supervised-learning BERT models in identifying documented goals-of-care discussions for hospitalized patients with serious illness.

| Dataset | NLP model (machine learning approach) | AUC | AUPRC | Maximal $F_1$ |
| --- | --- | --- | --- | --- |
| *Note-level classification* |  |  |  |  |
| Trial 1 test set (n=2,974) | Llama 3.3 (zero-shot ML) | 0.961 | 0.795 | 0.74 |
|  | BERT (supervised ML) | 0.962 | 0.789 | 0.74 |
| Trial 2 held-out test set (sample of n=2,136)<sup>a</sup> | Llama 3.3 (zero-shot ML) | 0.979 | 0.873 | 0.83 |
|  | BERT (supervised ML) | 0.981 | 0.874 | 0.83 |
| *Patient-level classification over ~30-day period* |  |  |  |  |
| Trial 1 test set (n=160) | Llama 3.3 (zero-shot ML) | 0.912 | 0.885 | 0.80 |
|  | BERT (supervised ML) | 0.898 | 0.833 | 0.80 |
| Trial 2 held-out test set (n=617)<sup>a</sup> | Llama 3.3 (zero-shot ML) | 0.977 | 0.955 | 0.89 |
|  | BERT (supervised ML) | 0.981 | 0.952 | 0.89 |

Abbreviations: AUC, area under the receiver operating characteristic [ROC] curve; AUPRC, area under the precision-recall curve; BERT, Bidirectional Encoder Representations from Transformers; ML, machine learning; NLP, natural language processing. The $F_1$ score is defined as the harmonic mean of positive predictive value (precision) and sensitivity (recall); the maximal $F_1$ is the highest observed $F_1$ across all discrimination thresholds.
<sup>a</sup> The Trial 2 held-out test set contained 11,574 notes, of which 2,136 with known true values were sampled for evaluation of note-level performance. Patient-level performance was evaluated by comparing NLP analysis of all 11,574 notes against the NLP-screened human-abstracted gold standard for all 617 patients.

**Figure 1.**
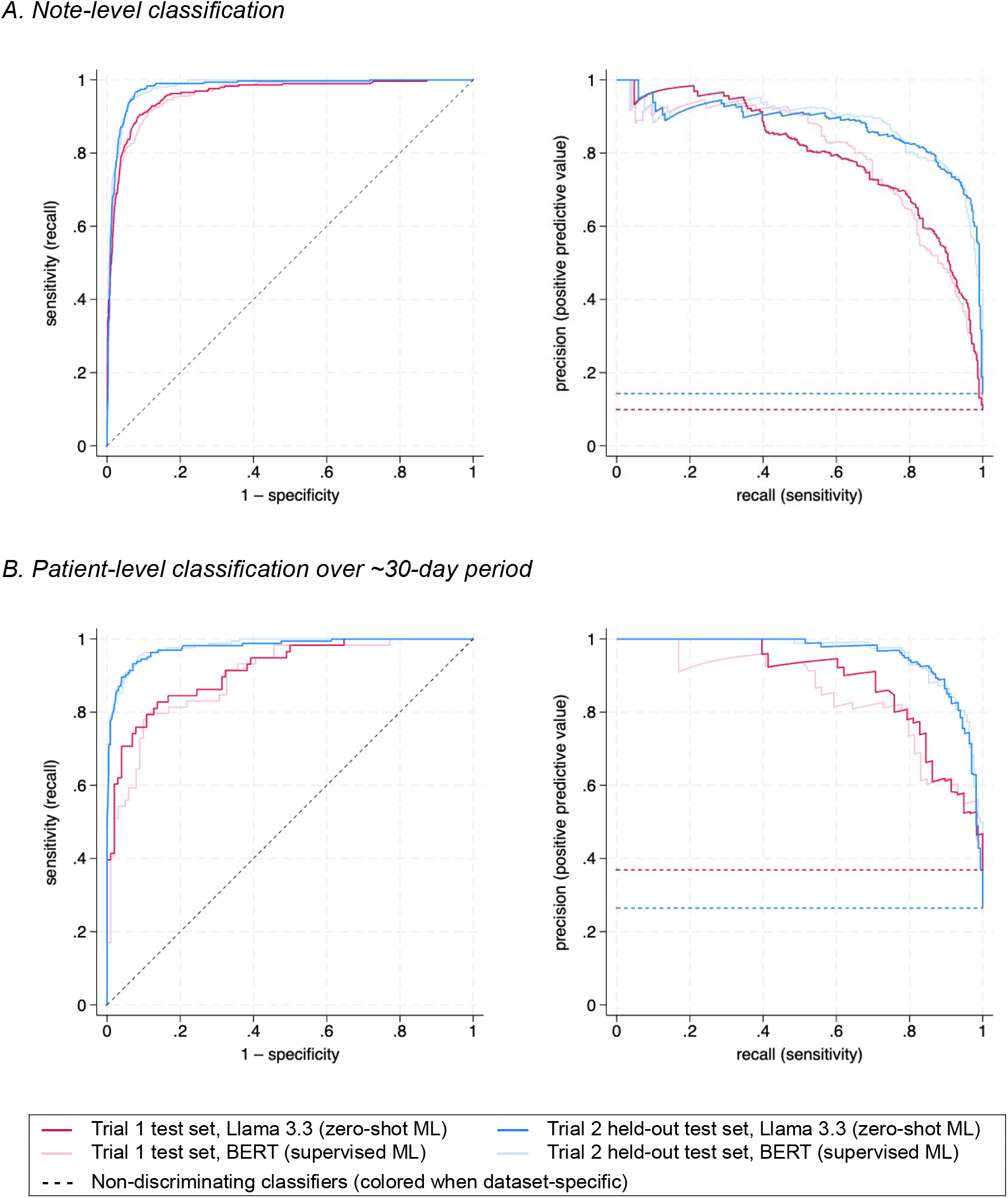
Performance of zero-shot Llama 3.3 vs. supervised-learning BERT models in detecting documented goals-of-care discussions for hospitalized patients with serious illness.

Notably, we observed higher performance in the Trial 2 held-out test set for both Llama and BERT models than in the Trial 1 test set. The difference in performance between datasets (**Figure 1**, red vs. blue lines) exceeded differences in performance between models (**Figure 1**, dark vs. light lines).

## Discussion

In this report, we demonstrated that a zero-shot LLM with no task-specific training can achieve comparable performance to a task-specific supervised learning BERT model trained on thousands of manually labeled EHR notes to identify documented goals-of-care discussions.

Although it hardly seems noteworthy that a 70-billion-parameter LLM can achieve comparable performance to a 110-million parameter BERT model, the ability to measure linguistically complex outcomes using a zero-shot prompting strategy—which is only possible with LLMs—is of great practical importance to clinical researchers. The training data alone for the referent BERT model was acquired over the course of 287 human abstractor-hours over a 4-month period—a ∼$20,000 investment to measure a single linguistic construct that must be repeated for any related constructs of interest, or even for different operational definitions of the same construct.^19^ This cost barrier poses significant challenges for researchers interested in measuring multidimensional constructs for which definitions are likely to differ appropriately from study to study.^37,38,35^ In contrast, iterative development of an LLM prompt is a faster process that may be quickly replicated or adapted to different constructs, allowing investigators to pivot toward different measures and outcomes between studies while incurring minimal repeated costs. This is especially important for linguistically complex outcomes such as goals-of-care discussions, which are difficult to measure using rule-based methods.^12^ The ability to identify linguistically complex constructs at scale also has implications for mixed-methods researchers, in that measures that were previously unquantifiable may be quantified.^39–42^

Despite their advantages, zero-shot methods have their disadvantages as well. First, zero-shot prompting relies heavily on the model’s pre-training from its developers. This dependency may introduce instability and irreproducibility as changes between model versions introduced by the developer may alter the model’s response to a given prompt in unpredictable ways. Second, LLMs are computationally resource-intensive, often requiring upgraded hardware to deploy at scale. The inefficiency of these models may limit their use cases, at least until LLM-capable hardware or institutionally-approved secure cloud environments for their use are widely available. Active efforts in the machine learning community to shrink and streamline LLMs may also help improve their efficiency.^43^ Third, LLMs suffer from a “black box” phenomenon, in which the reasons for a model’s predictions are opaque to their operators. However, this limitation— which is shared by nearly all ML models—is potentially easier to overcome with LLMs than with previous-generation models such as BERT, as prompting strategies that ask LLMs to follow specific patterns of logical reasoning have been shown to both be feasible to implement and also a means to generate more robust model output.^44^ LLMs also contain embedded biases that arise from their training data.^45^ However, prompting strategies that guide LLMs through cognitive processes are an intriguing potential solution to reduce biases in LLM responses, a strategy that is not available to previous-generation models.^46^

Performance of both models was similar within each dataset, but differed noticeably between the Trial 1 test set and the Trial 2 held-out test set (**Figure 1**). Although BERT was itself used to screen records for human ground-truth adjudication in the Trial 2 dataset, the screening threshold was exceedingly permissive (i.e., sensitive), and additionally, any such bias should not affect the performance of the zero-shot Llama 3.3 model. We suspect that observed differences in performance between datasets are probably attributable to differences between the datasets such as label quality and data curation procedures, differences in patient population, or secular trends in documentation. We believe this observation highlights the importance of evaluating NLP performance within specific use contexts, as both goals-of-care communication practices and their associated documentation practices are likely to differ between patient populations and clinical environments.

LLMs have many potential applications in serious illness communication. Beyond supporting clinical research, LLMs may assist clinicians by reviewing prior goals-of-care discussions, summarizing illness trajectories, identifying patients who may need serious illness conversations, supporting documentation, and enabling quality monitoring. Looking ahead, more advanced models with patient-specific inputs could even serve as adjunctive communication tools tailored to individual patients. However, realizing these potential applications will require overcoming both technical and implementation barriers. On the technical side, LLMs demand computational resources that often exceed current clinical hardware, making real-time chart-wide analysis impractical at the point of care. These challenges are likely surmountable through deeper EHR-cloud integration, advances in model efficiency, and adaptive AI systems that route tasks between lightweight and resource-intensive models. In contrast, implementation barriers will be far harder to address. Clinical workflows are already burdened by high cognitive load, and serious illness communication is further strained by high stakes, strong emotions, deficits in trust and trustworthiness, inequities in care delivery, and resistance to impersonal technologies. Successfully applying LLMs in this context will therefore require disciplined, methodical development that includes patient- and clinician-centered design, robust attention to safety and equity, and meaningful engagement of all stakeholders—practices that are too often overlooked in EHR development. The *potential* of LLMs in this domain is very real, but translating promises into practice hinges less on computational power and more on human factors, workflow design, and stakeholder trust.

Our study has important limitations. First, we evaluated a single operational definition of a single linguistically complex outcome. While much of the promise of this work lies in measuring related constructs, our findings and implications may not apply to all linguistic constructs. Even within the community of investigators interested in measuring goals-of-care content, other operational definitions of the same construct are more amenable to measurement using rule-based and other methods.^15,16,20^ Differences in the performance of rule-based methods across these studies^12,20^ illustrates how minor differences in operational definitions (e.g., the inclusion vs. exclusion of routine code status discussions) can lead to important differences in the choice of best measurement tools. Second, all data were collected from a series of randomized trials of related interventions performed at a single multi-hospital health system in Washington State, which limits generalizability outside of this setting. Although this study does not account for the trial interventions, our understanding of the interventions (which are all low-touch communication prompting interventions), as well as qualitative work in progress, suggest that the interventions had very little systematic effect on the location or linguistic structure of clinicians’ goals-of-care documentation. Third, our held-out test set was incompletely adjudicated by humans due to its sheer size. However, we feel that the high expected sensitivity of our adjudication method for the held-out test set lends advantages in generalizability that far outweigh limitations in fidelity. This tension between scale and fidelity has long been known to the NLP community, and we anticipate that navigating this tension will continue to be an area of great interest in the intersection of NLP and epidemiology.^47–49^

## Conclusion

In measuring documented goals-of-care discussions for hospitalized adults with serious illness, a zero-shot Llama 3.3 large language model with no task-specific training achieved comparable classification performance to a task-specific supervised-learning BERT model that had been trained on thousands of manually labeled EHR notes. The advent of large language models and their facilitation of zero-shot methods have important implications for clinical and health services researchers interested in leveraging the EHR to measure linguistically complex exposures and outcomes.

## Data Availability

Due to the identifiable nature of the protected health information evaluated in this study, freetext data are not available for disclosure without institutional approval and authorization.

## Author contributions

Dr. Lee had full access to all of the data in the study and takes responsibility for the integrity of the data and the accuracy of the data analysis.

## Acknowledgement

The authors are grateful for the contributions of the late J. Randall Curtis, MD, MPH, who was a founding principal investigator of this research program.

## Appendix 1. Prompt and prompt development for zero-shot Llama 3.3 language model

### Prompt development

Our goal was to develop a zero-shot LLM prompt to measure an outcome previously annotated by human abstractors in our pilot and test sets (codebook published in Lee RY et al, *JAMA Network Open* 2023;6(3):e231204, Supplement 1, eAppendix 2). Beginning with a simple, expert-composed prompt, the authors met weekly over 9 months to qualitatively review LLM outputs on exemplar pilot-set records. Prompt iterations prompted for both binary adjudications and machine-generated reasoning. Based on qualitative review of false positives and negatives and published approaches to prompt engineering, we proposed and tested prompt revisions designed to improve alignment between the LLM’s apparent interpretation of the task and the outcome definition used for annotation. Exemplar records were initially sampled randomly across positive and negative strata; and, in later runs, enriched for edge cases to probe specific topical misunderstandings.

This iterative process produced a definition-informed prompt that defined key concepts and instructed the LLM to identify a composite outcome comprising three constructs: goals-of-care discussions (adapted from Secunda et al, *J Gen Int Med* 2023;35(5):1559-1566), documentation about advance care planning documents, and designation of surrogate decision-makers. The prompt also specifically excluded code-status-only discussions. Once the core definitional content of the prompt was finalized, instructions to generate reasoning were replaced with requests for a binary (“YES/NO”) adjudication and an “impact score.” Although the LLM was asked to consider the impact score in forming judgements, it was not asked to (and did not) output a numeric value. The final prompt was then quantitatively evaluated on the Trial 1 test set and validated on the Trial 2 held-out test set, as described in this report.

### Prompt text

~~~
Before we begin, let’s define a few key concepts:
- GOALS-OF-CARE DISCUSSIONS are defined as discussions between clinicians and patients/families about the patient’s aims of medical care, which are ideally informed by their underlying values and priorities, established within the current clinical context, and used to guide decisions about the use of or limitations on life-sustaining treatments.
- CODE STATUS refers to inpatient orders for a patient that specify what is to be done in the event of cardiac or respiratory arrest--i.e., whether to perform CPR or not (“DNR”), whether to intubate or not (“DNI”). Code status discussions are one type of goals-of-care discussion, albeit one of narrow scope.
- “Advance care planning documents” are structured documents such as advance directives and POLST forms that may provide insight into a patient’s goals of care or treatment preferences for future health states, as well as durable power of attorney (DPOA) forms. One component of advance care planning, the designation of a “surrogate decision-maker” or “durable power of attorney”, refers to designation by a patient of a proxy who can make healthcare-related decisions on their behalf if they are incapacitated.
Our research outcome, COMPOSITE_JS, is defined as ANY of three constructs: (1) goals-of-care discussions that extend -beyond-decisions relating to code status alone; (2) incident designation or review of surrogate decision-maker or durable power of attorney (not just citations of past documentation); and, (3) incident completion or review of advance care planning documents such as advance directives or POLSTs (not just citation of prior documents). COMPOSITE_JS should -not-include text that only references statements aspiring to future goals-of-care discussions, treatment-specific therapeutic goals, or documentation focused on discharge or disposition planning (e.g., plans for post-discharge placement).
Using the medical record excerpts provided in INPUT, do the excerpts in INPUT reflect documentation of a discussion meeting criteria for COMPOSITE_JS? Consider the evidence and assign an impact score adding up to 10, where higher scores indicate stronger influence on the final decision. If key elements are missing (e.g., no documented goals-of-care discussion, no incident designation/review of a surrogate, or no review/completion of advance care planning documents), assign higher scores to reflect their critical absence.
Please only answer YES or NO.
Use the following example format:
ANSWER: [YES/NO]
INPUT = [*insert candidate electronic health record excerpt*]
~~~

